# Pitfalls in pre-operative prediction of lymph node metastasis in early endometrial cancer

**DOI:** 10.1101/2020.10.21.20216622

**Authors:** Pampapati Veena, Rajalakshmi Ilango, Jayalakshmi Durairaj

## Abstract

**Objective:** The role of lymphadenectomy in early stage endometrial cancer is controversial as it is associated with intra-operative complications and its therapeutic benefit is not established. Prediction of lymph nodal metastasis so as to perform selective lymph node dissection is desirable. This study was conducted to study grade of the tumor obtained by endometrial biopsy specimen and depth of myometrial invasion assessed by imaging pre-operatively as predictors of lymph nodal metastasis in early endometrial cancers.

**Methods:** Our study was a cross sectional study done in a tertiary care center in south India, where 100 patients were studied from August 2016 to May 2018. After Ethical Committee clearance, all patients who were diagnosed with early endometrial cancer and who underwent surgery in our hospital were included in the study after getting informed consent. Pre-operative tumor grade and depth of myometrial invasion were studied as predictors of lymph nodal metastasis. They were also correlated with post-operative grade and myometrial invasion.

**Results:** The present study recruited 100 women of which 3 cases were excluded because of non-endometrioid histology. The incidence of positive lymph node metastasis in our study was 18.6%. Both pre-operative tumor grade and depth of myometrial invasion were not significantly associated with lymph node metastasis. There was significant variation between pre-operative and post-operative tumor grade and depth of myometrial invasion. Among post-operative histopathological factors, only lymphovascular space invasion was found to be significantly associated with lymph node metastasis.

**Conclusion:** In our study, neither pre-operative nor post-operative grade of the tumor and depth of myometrial invasion were significantly associated with lymph node metastasis. There was considerable variation between pre-op and post-op grade of the tumor making pre-op grade an unreliable factor in predicting lymph node metastasis in endometrial cancer. Among post-operative histopathological factors, only lymphovascular space invasion was found to be significantly associated with lymph node metastasis.

**Key message:** Considerable variation between pre-op and post-op grade of the tumor makes pre-op grade an unreliable factor in predicting lymph node metastasis in endometrial cancer

## INTRODUCTION

The role of lymphadenectomy in early stage endometrial cancer is controversial. Several retrospective studies have suggested that there is therapeutic benefit with lymphadenectomy and recommended that routine pelvic lymphadenectomy should be performed in low risk endometrial cancer patients in addition to total abdominal hysterectomy and bilateral salpingo-oophorectomy and para-aortic lymphadenectomy can be performed only in patients with high risk characteristics.^1-3^ However, many prospective trials done recently found that there was no significant improvement in the disease free survival or overall survival with lymphadenectomy.^4, 5^ Moreover, retro-peritoneal lymphadenectomy is associated with intra-operative complications like injury to the major vessels, nerves, adjacent viscera and post-operative complications like wound infection, lymphedema, lymph-cyst formation, chronic pain etc.^4, 5^ Due to these reasons, lymphadenectomy in these patients is still controversial with varied practice among surgeons^6^. Therefore, it becomes very important to identify patients who will benefit from lymphadenectomy. Numerous studies have been conducted to find out the important predictive factors that can guide us to identify the patients who are at high risk of developing nodal metastasis and who will benefit from retroperitoneal lymphadenectomy.^7, 8, 9^ To consider grade of the tumor as a factor predicting lymph node involvement, significant variation in up to 30% of cases has been noted between pre-operative grade and post-operative grade.^10^ This study was conducted to study grade of the tumor obtained by endometrial biopsy specimen and depth of myometrial invasion assessed by imaging pre-operatively as predictors of lymph nodal metastasis in early endometrial cancers

## MATERIALS AND METHODS

Our study was a cross sectional study done in a tertiary care center in south India, where 100 patients were studied from August 2016 to May 2018. After Ethical Committee clearance, all patients diagnosed with early endometrial cancer and underwent surgery in our hospital were included in the study after getting informed consent. Patients who were not fit for surgical management and those who were not willing to participate in the study were excluded.

### Sample size calculation

Using the values of overall incidence of lymph node metastasis in previous studies and with a power of 80% and alpha error of 5% in open epi and considering that approximately 25% of patients would be in the high risk group, the sample size was calculated to be 69 patients in low risk group and 23 patients in high risk group. To get a sample size of 23, 92 subjects with endometrial cancer need to be included in the study. Hence the sample size was estimated to be 100 to account for the patients lost to follow up.

### Procedure

All the eligible patients were counselled and explained about the study protocol. A written informed consent was obtained from eligible study subjects before recruitment, both in English and local language known to the study subjects. Relevant socio demographic details, age, obstetric/past/family history were collected. Histology and pre-operative grade were assessed by HPE of endometrial biopsy. After the resection of the specimen, 10 ml of formalin was injected into the uterine cavity to preserve the endometrial tissue. Histology and grade of tumor, depth of myometrial invasion, cervical stromal invasion, lymphovascular space invasion and evidence of lymph node involvement was assessed post-operatively by HPE of the operated specimen. Myometrial invasion was measured in millimeters from endo-myometrial junction and the correlation between depth of myometrial invasion with lymph node metastasis was assessed. Also, the correlation between pre-operative histo-pathological factors, tumor grade, myometrial invasion and lymphovascular space invasion to lymph node metastasis was evaluated. And the intra-operative and immediate post-operative complications of lymphadenectomy were noted. Data was collected in a pre-designed proforma from all the women undergoing surgery for early endometrial cancer in JIPMER in the proposed duration during the pre-operative, intra-operative and post-operative stay in the hospital. Patients were followed up for a period of 6 weeks and HPE reports were reviewed and the correlation between grade, depth of myometrial invasion and LVSI with lymph node metastasis in early endometrial cancer were analyzed.

### Analysis

Patient characteristics such as age, BMI, parity, socioeconomic status, menopausal status, histology type of cancer, pre/post-operative grade tumor, myometrial invasion, lympho-vascular invasion, complications due to surgery were summarized as frequency and percentage. Association between myometrial invasion, lymphovascular invasion, pre/post-operative grade and lymph node involvement was studied using odds ratio and 95% CI. Chi-square test was applied to assess for statistical significance. Correlation between pre-operative and post-operative grade of tumor was assessed using percentage agreement. Association between independent variables and dependent variable (lymph node involvement) was studied using univariate analysis and later with multivariate logistic regression. p value <0.05 was set as significance level and analysis was done using SPSS version 19.0 software.

### Ethical approval

Institute Post-graduate Research Monitoring Committee Certificate no. 18.04.16/15 dated 8^th^ May 2016 and Institute Ethics Committee Certificate no. JIP/IEC/SC/2016/30/962 dated 4^th^ of August 2016. Informed written consent was obtained from all participants.

## RESULTS

Present study recruited 100 women of which 3 cases were excluded because of non-endometrioid histology. Regarding the distribution of socio-demographic and other baseline characteristics of the patients included in our study, most of the patients were multi-parous (88.6%) and postmenopausal (84.5%) and most of them were obese with a mean BMI of 31.92 ± 2.72 SD. The mean age at menarche was found to be 14.73 years ± 1.25 SD and mean age at menopause was found to be 49.27 years ± 3.09 SD. Regarding the distribution of medical co-morbidities, diabetes mellitus and hypertension accounted for 55.7 % and 44.5% of the study population respectively.

Pre-operatively, histology and grade of the tumor was assessed in the endometrial biopsy specimen and depth of myometrial invasion was assessed by pelvic USG/MRI for all patients. Majority of the patients in our study (83.5%) had grade 1 disease and 14 (14.4%) and 2 (2.1%) patients had grade 2 & 3 endometrioid adenocarcinoma respectively. Depth of myometrial invasion was assessed as <50% or >50%, and almost equal numbers belonged to both the groups.

All patients in our study underwent extra-fascial hysterectomy with pelvic lymphadenectomy with or without bilateral salpingo-oophorectomy. Post-operatively, the resected surgical specimen was subjected to histo-pathological examination and grade of the tumor, depth of myometrial invasion, presence or absence of cervical stromal invasion, presence or absence of lymphovascular space invasion, presence or absence of lymph node metastasis, final stage of the disease and any post-operative complications were noted. Although patients with endometrial cancer confined to the uterus pre-operatively were included in our study, 23.7% got upstaged after histo-pathological examination of resected specimen; 5.1% showed cervical stromal invasion (stage 2) and 18.6% of the patients got upgraded to stage 3 post-operatively. Majority of the patients had grade 1 disease (67%) and grade 2 and grade 3 disease accounted for 26.8% and 6.2% respectively. Regarding myometrial invasion, 36.1% had <50% and 63.9% had >50% myometrial invasion. Seventeen patients showed positive lymphovascular space invasion accounting for 17.5%. Among the 97 patients included in the study, 18 (18.6%) patients had positive lymph node metastasis and lymph node metastasis was absent in 79 (81.4%) patients.

Out of 97 patients included in the study, 26 patients (26.8%) experienced complications; 4 (4.1%) were intra operative and 22 (22.68%) were post-operative complications. All the intra-operative complications were due to vascular injury and none of the patients had injury to the viscera. Among the post-operative complications, majority were wound infections and most of the wound infections occurred among diabetic patients. Other complications observed were urinary retention and complete wound dehiscence. One mortality was encountered in our study which was due to complete wound dehiscence followed by post-operative sepsis and dyselectrolytemia culminating in sudden cardiac arrest due to pulmonary embolism.

In our study, based on the final histopathological examination report and the presence or absence of high-risk factors, patients were subjected to either radiotherapy alone or sequential chemotherapy followed by radiotherapy. Totally, 41 patients (42.2%) received adjuvant therapy in our study; 23 - radiotherapy alone and 18 - sequential chemotherapy followed by radiotherapy.

### Correlation between pre-operative grade and post-operative grade (percentage agreement)

Among 81 cases pre-operatively diagnosed as grade 1, only 70.4% remained as grade 1 and rest were upgraded to grade 2 (27.2%) and 3 (2.5%) post-operatively. Among 14 cases of grade 2 diagnosed pre-operatively, only 28.6% remained grade 2, 50% were downgraded to stage 1 and 21.4% patients got upgraded to grade 3 post-operatively and among 2 cases of grade 3 tumor diagnosed pre-operatively, 1 (50%) case remained as grade 1 and 1 (50%) downgraded to grade 1 (Table 1).

**Table 1:** Pre-operative grade versus post-operative grade

| Pre-operative grade | Post-operative grade |  |  | Total | Significance |
| --- | --- | --- | --- | --- | --- |
|  | Grade 1<br>N (%) | Grade 2<br>N (%) | Grade 3<br>N (%) |  |  |
| Grade 1 | 57 (70.4%) | 22 (27.2%) | 2 (2.5%) | 81 (100%) | 0.21* |
| Grade 2 | 7 (50%) | 4 (28.6%) | 3 (21.4%) | 14 (100%) |  |
| Grade 3 | 1 (50%) | 0 (0%) | 1 (50%) | 2 (100%) |  |
| Total | 65 (67%) | 26 (26.8%) | 6 (6.2%) | 97 (100%) |  |

\*p value=0.21 by Kappa statistics

### Association between pre-operative grade and myometrial invasion with pelvic lymph node metastasis

In the pre-operative assessment, only 2 patients out of 97 had grade 3 disease. The association between pre-operative grade of the tumor and lymph node metastasis was not statistically significant in our study (p=0.338). Though the proportion of patients with positive pelvic lymph node metastasis was found to be higher among patients with deep myometrial invasion, the association did not reach statistical significance (p value=0.435) (Table 2).

**Table 2:**
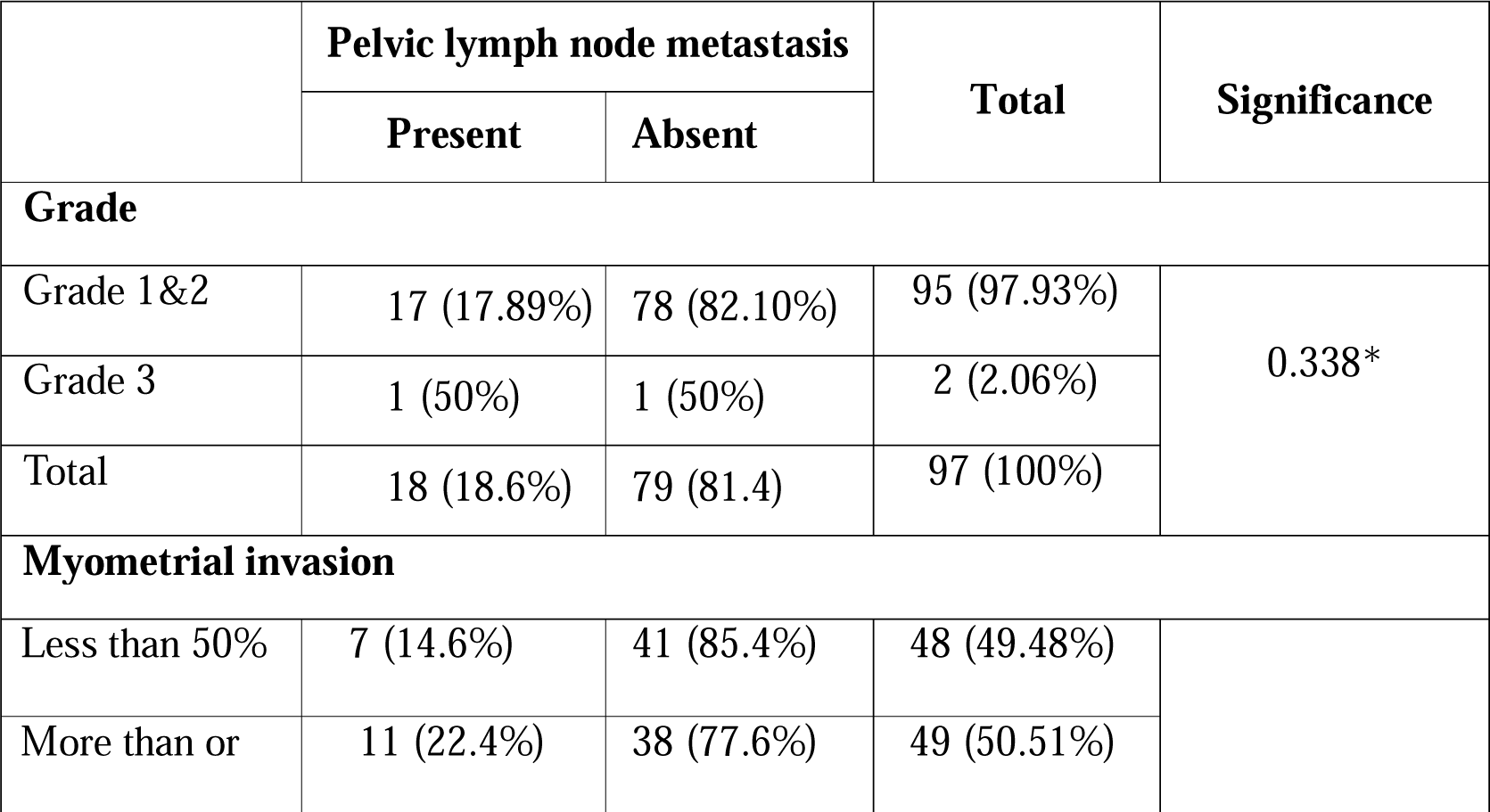
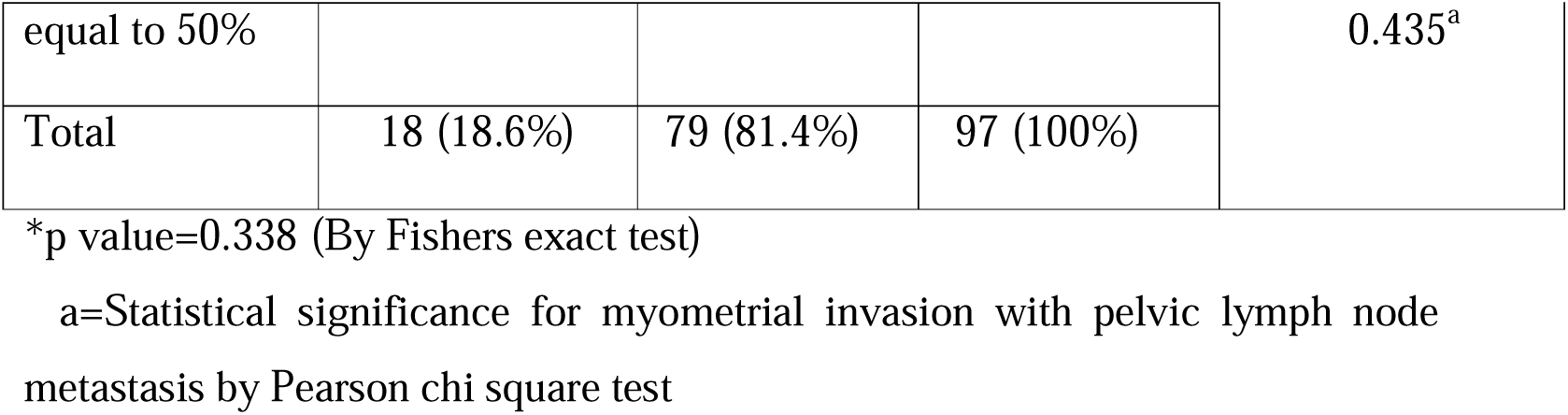
Association between pre-operative grade and myometrial invasion with pelvic lymph node metastasis

### Association between post-operative grade, myometrial invasion and lymphovascular space invasion with pelvic lymph node metastasis

In our study, pre-operative grade did not show any significant association with lymph node metastasis. As there was considerable variation between pre-operative grade and post-operative grade in our study, the association between post-operative grade and lymph node metastasis was analyzed separately. Fifty percent of patients with grade 3 disease had lymph node metastasis when compared to 17.5% of grade 1 and 2 disease. Although this difference was not significant statistically, the p value of the association between pre-operative grade and lymph node metastasis and post-operative grade and lymph node metastasis was different in our study (0.338 vs 0.08). (Tables 2 and 3). Among 97 patients in the study, lymphovascular space invasion was present in 17 (17.5%) patients and 82% of these patients had lymph node metastasis when compared to 17.6% among patients with no lymphovascular space invasion as shown in table 3. This difference was statistically significant with a p value of <0.001. The association between depth of myometrial invasion and lymph node metastasis was not found to be statistically significant (p value – 0.133) in our study, the details of which is shown in table 3.

**Table 3:** Post-operative grade and pelvic lymph node metastasis

|  | Pelvic lymph node metastasis |  | Total | Significance |
| --- | --- | --- | --- | --- |
|  | Present | Absent |  |  |
| Post-operative grade |  |  |  |  |
| Grade1 | 10 (15.3%) | 55 (84.61%) | 65 (100%) | 0.08* |
| Grade 2 | 5 (19.23%) | 21 (80.76%) | 26 (100%) |  |
| Grade 3 | 3 (50%) | 3 (50%) | 6 (100%) |  |
| Total | 18 (18.55%) | 79 (81.44%) | 97 (100%) |  |
| LVSI |  |  |  |  |
| Present | 14 (82.4%) | 3 (17.6%) | 17 (17.52%) | <0.001<sup>a</sup> |
| Absent | 4 (5%) | 76 (95%) | 80 (82.47%) |  |
| Total | 18 (18.6%) | 79 (81.4) | 97 (100%) |  |
| Myometrial invasion |  |  |  |  |
| <50% | 5 (14.28%) | 30 (85.71%) | 35 (100%) | 0.133<sup>b</sup> |
| >50% | 13 (21%) | 49 (79%) | 62 (100%) |  |
| Total | 18 (18.55%) | 79 (81.45%) | 97 (100%) |  |

\*p value = 0.08 (by linear association)
a- Statistical significance for LVSI with pelvic lymph node metastasis by Fishers Exact test
b- Statistical significance for myometrial invasion with pelvic lymph node metastasis by linear by linear association

We further divided depth of myometrial invasion as less than 25%, 25-50%, 50-75%, 75%-100% to assess the association with lymph node metastasis. There were no lymph nodal metastases among women with <25% myometrial invasion and almost 1/3rd of patients with >75% myometrial invasion had lymph node metastasis (Table 4). But these differences did not reach statistical significance.

**Table 4:** Depth of myometrial invasion and lymph node metastasis

| Myometrial invasion<br>in percentage | Pelvic lymph node<br>metastasis |  | Total | Significance<br>(p value) |
| --- | --- | --- | --- | --- |
|  | Present | Absent |  |  |
| <25% | 0 (0%) | 6 (100%) | 6 (100%) | 0.133<sup>a</sup> |
| 25-50% | 5 (17.2%) | 24 (82.8%) | 29 (100%) |  |
| 50-75% | 6 (15.4%) | 33 (84.6%) | 39 (100%) |  |
| 75-100% | 7 (30.4%) | 16 (69.6%) | 23 (100%) |  |
| **Total** | 18 (18.6%) | 79 (81.4%) | 97 (100%) |  |

a- Statistical significance for myometrial invasion with pelvic lymph node metastasis by linear by linear association

We had also assessed the depth of myometrial invasion in millimeters in an attempt to find out depth of invasion in millimeters above which there is a significant increase in incidence of pelvic lymph node metastasis. ROC curve was used for determining the significant depth of myometrial invasion associated with pelvic lymph node metastasis. The area under the curve was observed to be 0.6 and 95% confidence interval of 0.49 to 0.77. The cut-off value for depth of myometrial invasion above which there was increased incidence of lymph node metastasis was found to be 17 mm with a sensitivity of 66% and specificity of 55%. The incidence of lymph node metastasis in patients with myometrial invasion less than 17 mm and more than or equal to 17 mm is shown in table 5.

**Table 5:** Depth of myometrial invasion in millimeters and lymph node metastasis

| Depth of<br>myometrial<br>invasion in<br>mm | Pelvic lymph node<br>metastasis |  | Total | Significance<br>(p value) |
| --- | --- | --- | --- | --- |
|  | Present | Absent |  |  |
| **Less than 17 mm** | 6 (12.2%) | 43 (87.7%) | 49 (50.51%) | 0.48<sup>a</sup> |
| **More than or equal**<br>**to 17 mm** | 12 (25.0%) | 36 (75.0%) | 48 (49.48%) |  |
| **Total** | 18 (18.6%) | 79 (81.4%) | 97 (100%) |  |

a- Statistical significance for myometrial invasion in mm with pelvic lymph node metastasis by Fisher's exact test

DISCUSSION

Endometrial cancer usually presents in early stage and deciding the extent of surgery is very important for optimal management of these women. Pelvic and para-aortic lymphadenectomy is associated with significant complications and its therapeutic benefit is still controversial. We also encounter women who have undergone hysterectomy without staging for abnormal uterine bleeding and later found to have endometrial carcinoma in histo-pathological examination of the resected specimen. Sometimes it is difficult to convince these women for repeat surgery for the purpose of staging and it is important to assess the association of various histopathological factors in the hysterectomy specimen with lymph nodal metastasis to decide about the adjuvant therapy. With this background, we studied 97 women with apparently early stage endometroid type of endometrial carcinoma to assess the association of various histopathological factors and imaging findings with lymph nodal metastasis. Grade of the tumor assessed in the endometrial biopsy specimen and depth of myometrial invasion determined by imaging are the two tools currently available to decide the extent of surgery. Regarding grade as a predictive factor for nodal metastasis, in our study pre-operative grade did not show a significant association with metastatic nodal disease in univariate analysis (p value-0.338). There are conflicting reports in the literature regarding grade as the predictor of lymph nodal metastasis. A study by Toptas et al.^11^ reported similar results as our study whereas two other studies had contradicting reports. Mariani et al.^12^ reported that tumor grade was significant factor that could predict nodal involvement in endometrial carcinoma. Another study which contradicted with our study was the study conducted by Rathod et al. which showed that the incidence of nodal disease was significantly higher in grade 3 and undifferentiated tumors than grade1 and 2 tumors. But, in their study non-endometrioid histology tumors were also included.^13^ In our study only 2 (2.1%) patients had pre-operative grade 3 tumors which could have significantly affected the association between grade and lymph node metastasis and with such a low number of patients with grade 3 disease the results could not be validated. In the study conducted by Mariani et al. which showed that there was a significant association between pre-operative grade of the tumor and lymph node metastasis, 28% of the study patients had grade 3 disease compared to 2.1% in our study.^12^

Moreover, in our study, there is significant variation between pre-op and post-op grading of tumor which further makes it difficult to use pre-op grade of tumor as a predictor of lymph nodal metastasis. Our study results are in association with another study conducted by Mahdi et al. in which nearly 9.7% of patients got upgraded to grade 3 from grade 1 post-operatively.^14^ In another study conducted by Neubauer et al. among patients with pre-operative grade 1 tumors, 21.1% patients were upgraded to grade 2 and 1.4% patients to grade 3 disease.^15^ In the study conducted by Son et al. preoperative tumor grade was upgraded in 5.6% of patients.^16^ In a retrospective study conducted by Rathod et al. in India, nearly 18.6% patients with grade 1 tumors preoperatively got upgraded to grade 2 tumors and 9.7% patients were upgraded to grade 3 tumors in post-operative specimen. They also noted that among that among 9.7% patients who were upgraded to grade 3 tumors in post-operative specimen, 50% had positive pelvic lymph node involvement. So, the authors concluded that there was a significant difference between pre-operative grade and post-operative grade of the tumor and pre-operative grade cannot be relied upon to decide the need for lymphadenectomy.^13^

Another pre-op predictor of lymph node metastasis, depth of myometrial infiltration suffers similar conflicting results in literature. In our study, pre-operatively determined depth of myometrial invasion was not found to be associated with nodal metastasis. Studies conducted by Son et al. and Toptas et al. also showed similar results that myometrial invasion was not a significant factor predicting nodal involvement.^11, 16^ Our study results were contradicted by the study done by Mariani et al. and Kamura et al. who had reported that depth of tumor infiltration into the myometrium was found to be independent factors associated with lymph node involvement.^12,17^

Among the post-op histopathological factors such as grade of tumor, depth of myometrial invasion and lymphovascular space invasion, lymphovascular space invasion remained as the only important factor predicting lymph node metastasis in our study. In our study, the pelvic lymph node metastasis was present in 18 (18.6%) patients among which 14 (82.4%) patients showed positive LVSI. Unfortunately, lymphovascular space invasion cannot be assessed preoperatively to aid in planning the extent of surgery. But among women with post-hysterectomy diagnosis of endometrial cancer, LVSI can be used to decide about the need for adjuvant therapy. Literature is rife with studies supporting this observation and multiple researchers have proved that LVSI is an independent factor for predicting lymph node involvement.^18-22^

Regarding the complications of lymphadenectomy in our study, incidence of intra-op complications was found to be 4.1% which was almost similar to that of the intra-op complication rates observed in the study conducted by Walker et al. in which the incidence of intra-op complications was found to be 4%.^23^ And regarding the post-op complications rates it was found to be 22.7% in our study which was almost similar to the study conducted by Franchi et al. in which the incidence of post-op complication was found to be 26.7%.^24^

## CONCLUSION

In our study, even among patients who had grade 1 and stage 1a disease pre-operatively, who were considered to be at low risk for nodal metastasis, 18.5% patients had positive lymph node involvement. Without lymphadenectomy these 18.5% patients would have been easily missed and down staged as stage 1 disease. And positive lymph node involvement was present in 14.6% of patients who had less than 50% myometrial invasion pre-operatively which again showed that even patients with <50% myometrial invasion are at significant risk of having a nodal disease. Considering the fact that patients with positive nodal metastasis have an option of curative post-operative adjuvant therapy and an excellent disease free survival and decreased morbidity and mortality rates associated with lymphadenectomy in the hands of experienced surgeon, we recommend that performing lymphadenectomy and detecting lymph node metastasis is comparatively better than omitting lymphadenectomy and missing out on lymph node metastasis. In apparently low risk patients, sentinel lymph mode sampling is a good alternative. Based on our results, we recommend assessment of lymph nodes for all endometrial carcinoma patients since there is a considerable variation between preoperative and post-operative grade on final histo-pathological examination.

### Copyright transfer statement

“I Pampapati Veena, the Corresponding Author of this article contained within the original manuscript which includes any diagrams & photographs and any related or standalone film submitted (the Contribution”) has the right to grant on behalf of all authors and does grant on behalf of all authors, a license to the BMJ Publishing Group Ltd and its licensees, to permit this Contribution (if accepted) to be published in any BMJ products and to exploit all subsidiary rights.

## Data Availability

Data will be available at request by e mail

## Authors’ contribution

Pampapati Veena: Protocol/project development, Manuscript writing/editing Rajalakshmi Ilango: Data collection or management, Data analysis Jayalakshmi Durairaj: Concept, Manuscript editing

